# Experiences of contraceptive use and contraceptive counseling among German psychology students: a cross-sectional online survey

**DOI:** 10.1101/2025.08.01.25332712

**Authors:** Johanna Seiwert, Anja Lindig

## Abstract

**Background:** In recent years, hormonal contraceptive use has declined in Germany, yet little is known about the contraceptive needs and preferences of its population. This study addresses this gap by (1) identifying attitudes, subjective knowledge, and information needs of psychology students regarding contraceptive methods, (2) evaluating person-centeredness (PC) and shared decision-making (SDM) in contraceptive counseling, and (3) comparing users of hormonal and non-hormonal methods regarding attitudes, knowledge, and age.

**Methods:** This cross-sectional study was conducted via an online survey among psychology students in Germany who were biologically capable of becoming pregnant. PC in counseling was assessed using the Person-Centered Contraceptive Counseling Scale (PCCC), while SDM was measured using the CollaboRATE questionnaire and Control Preference Scale. Attitudes, knowledge, and information needs were evaluated through adapted or self-developed items. The PCCC was translated and adapted using a team translation protocol (TRAPD).

**Results:** Of 126 participants, 82.6% used at least one contraceptive method, mainly condoms (43.8%) and the combined pill (19.8%). Hormonal users were younger (M=22.84) than non-hormonal users (M=25.40). Attitudes toward hormonal contraception were generally negative. Participants reported good or basic knowledge of 9 out of 15 methods; non-hormonal users knew more methods than hormonal users. Gynecologists (36.7%) were the most frequent information source. About 35.7% had received contraceptive counseling in the previous year but reported low satisfaction with the information provided. Overall, PC and SDM were only partially implemented in contraceptive counseling.

**Conclusion:** Findings highlight the need for more person-centered counseling to support informed decisions. The results can inform interventions aimed at improving contraceptive knowledge among users and enhancing PC and SDM practices among gynecologists.

## BACKGROUND

The global decline in the fertility rate, meaning the average number of births per woman, has become notable in recent decades (1). Whereas in 1960 there were globally five births per woman, the number has halved to 2.5 in 2023 (1). Alongside the prospective education and empowerment of women, the establishment of modern contraception in the 20th century was the main driving force behind this decline (1,2). As a result, women nowadays have more reproductive autonomy and options for family planning (1,3). Given the wide range of contraceptive methods available, it is essential for women to have access to reliable information about the various methods and their advantages and disadvantages (4). This allows them to choose a method that suits their individual lifestyle, family planning and risk factors best (4). Common considerations when choosing a contraceptive method include information about its effectiveness, potential side effects, convenience, impact on the menstrual cycle, and the influence on sexual satisfaction (5). Healthcare providers (HCPs) were shown to have the highest influence on women’s choice of contraceptive methods and recommend the oral contraceptive pill (OCP) and the male condom most often (6). However, recent data from a representative survey with 1.001 sexually active adults in Germany indicate a shift in contraceptive use: In 2023, 70% of participants used contraception (7). Among them, 53% relied on male condoms, while 38% used the OCP. This is an notable change from 2011, where 53% used the OCP and 37% used male condoms (7). Additionally, the use of intrauterine devices (IUDs; small, flexible T-shaped plastic device inserted into the uterus for contraceptive purposes) increased from 10% of the surveyed sexually active adults using contraception in 2011 to 14% in 2023 (7,8). As a reflection of the decrease in OCP use, women in Western countries have increasingly used various social media platforms over the past decade to express their dissatisfaction with OCPs and their decision to stop taking it, with the primary focus on their concerns about taking hormones in general (9). At the same time, there is a notable lack of knowledge regarding the mechanisms and effectiveness of different contraceptives (10–12). For instance, previous studies indicate that women often overestimate the effectiveness of OCPs while tending to undervalue the effectiveness of long-acting reversible contraception such as IUDs (4,10,11). This knowledge gap is particularly apparent among younger women, who generally have a lower level of contraceptive knowledge (13,14). Furthermore, subjective knowledge is reported to be lower among women who feel that they lack information about their contraceptive options, compared to those who feel sufficiently informed (15). Knowledge about contraceptive methods is shaped by personal and family experiences, the influence of the media and personal values (16–18). Therefore, the comprehension of information is a decisive factor in the successful utilization of contraception (14). A significantly higher level of knowledge and a more positive attitude towards the use of contraceptives was reported when patients have been informed by HCPs (19). This emphasizes the importance of HCPs providing high-quality information (20–22).

Fertility preferences are complex and there are various factors that can influence them, including social, religious aspects, family consent or medical factors (30). Preferences for contraceptive methods can also vary over a lifetime (30). Thereby, contraceptive counseling should take place in a patient-centered setting that enables informed decisions on contraceptive use (23–25). The concept of patient-centeredness (PC) comprises focusing on preferences, needs and values of patients ensuring the well-being of patients in terms of health promotion and maintenance (26). A central aspect of PC is shared decision-making (SDM) (27). SDM is defined as an interactive exchange between patient and HCP with the aim of reaching a responsible agreement based on the sharing of information and through the equal and active participation of both (28). Accordingly, the responsibility for selecting the appropriate treatment (or contraceptive method in our case) is shared between patient and HCP (29). SDM plays an important role in contraceptive counseling, as it helps patients to consider their preferences and values in relation to their contraceptive needs at different stages of their reproductive life (19,31). Contraceptive counseling based on PC and SDM can increase satisfaction, improve adherence and instructions for contraceptive methods, and promote contraceptive use (32). Thereby, the patient’s preferences regarding hormone use should be included in the decision-making process to maintain autonomy regard decisions on contraceptive use (33).

Apart from a few studies on contraceptive use, such as the ‘Thinking About Needs in Contraception’ (TANCO) study, there is little validated data on contraceptive needs and implementation of SDM and PC in contraceptive counseling among individuals in Germany (4).

Thus, the overall aim of this study was to understand differences between individuals using hormonal contraception (HC; HC-users) and those, not using HC (non-HC users), and their experiences in contraceptive counseling to shed light on possible reasons for the shift towards non-hormonal contraceptive methods and the declining number of HC users in Germany.

## METHODS

### Study aim

Therefore, the aim of the study was:

(1) to identify attitudes towards HC, subjective knowledge about contraceptive methods and information needs of psychology students requiring contraceptives;
(2) to assess PC and SDM in contraceptive counseling;
(3) and to compare HC users to non-HC users in terms of attitudes regarding HC, subjective knowledge about contraceptive methods and age.

### Study design

This study was a cross-sectional study applying a quantitative online survey. A between-subject design was employed. The study design and reporting followed the checklist for reporting results of the internet e-survey (CHERRIES) (see S1 Appendix, 34)).

### Participants

The study population included psychology students with a female anatomy. This included all individuals, who had a female reproductive system, independent from their contraceptive or sexual practices or identity. Henceforth, in this study the term “individuals with uterus” is used to describe the population of interest. Thereby, non-binary individuals and transmen affected by contraception needs were included as well. Furthermore, participants should be at least 18 years old and had to be enrolled in a psychology bachelor’s or master’s program at a German university or institution of higher education at the time of the survey. The necessary sample size was calculated a priori using the software G*Power (version 3.1.9.4) (35). This calculation was performed for a Student’s t-Test to detect a difference between two independent groups. Given an estimated effect size of 0.5, an α error probability of 0.05 and a statistical power of 0.95 for a one tailed analysis, the calculated sample size was 88 data sets per group, which means 176 participants in total. It could be expected that up to 20% have to be excluded because of incomplete data sets or not fulfilling the inclusion criteria. For this reason, the target sample size was determined to be 211 participants.

### Data collection

The link and QR code for the online survey were sent via e-mail to psychology faculties of all state universities and selected institutions of higher education in Germany that offered a bachelor’s or master’s degree in psychology at the time of the survey. The relevant faculty members, academic staff or student representatives were identified and contacted independently via internet research and asked to forward the survey to their students. Additionally, the link and QR code were distributed via social media platforms including Instagram, Facebook and WhatsApp. The survey was conducted in German using the LimeSurvey platform, participation was voluntary and not remunerated. Responses provided were automatically recorded and stored in the LimeSurvey database. Information on data collection, analysis and data management was provided at the beginning of the online survey, and participants were given the option of downloading the data management concept as a PDF. If willing to participate, participants were required to confirm their consent by selecting the respective option. A progress bar was employed to indicate the participant’s progress while answering the survey and a back button allowed them to navigate between pages at any time. Cookies were used to avoid duplicate participation. The first access was counted, regardless of the completeness of the answers to the questions. It was a priori defined that the online survey will be closed as soon as the calculated sample size has been reached or after 6 weeks. Data collection lasted from May 17^th^ 2024 to June 28^th^ 2024.

### Instruments

Validated questionnaires were used to assess SDM and PC (31,36–41). The Person-Centered Contraceptive Counseling scale (PCCC), as a validated scale measuring PC in contraceptive counseling, was translated into German using a team translation protocol (Translation, Review, Adjudication, Pretesting and Documentation (TRAPD)), conducted by AL and two other researcher of the research group (VL, MR, see List of Abbreviations) (31,36,42). For the SDM assessment, the three-item CollaboRATE scale in a validated German version was used (40). The degree of desired control in medical decisions was assessed with the one-item Control Preference Scale (CPS), which is used widely in a translated but not validated German version (40,41,43,44). Further survey items were used to assess demographic characteristics of participants, their contraceptive behavior, attitudes regarding HC, subjective knowledge about contraceptive methods and information needs regarding contraception. Most of them were extracted from various relevant studies and adapted and translated, if necessary, as shown in Table 1. Items regarding contraceptive behavior were self-developed. When a translation from English into German was necessary, this was carried out on an ad hoc basis and discussed by the study team (AL, JS) regarding comprehensibility.

**Table 1.** Details of measured constructs and assessed items.

| Construct | Description | Item number | Source | Adaption | Scale |
| --- | --- | --- | --- | --- | --- |
| **Attitude** | Personal opinions about hormonal contraception (HC) | 8 | 45,46 | Ad-hoc translated, adapted, change of scale from dichotomous to Likert scale | 5-point Likert scale from 1 "strongly disagree" to 5 "strongly agree" |
|  | Perceived side effects of HC | 1 | 45 | Ad-hoc translated, response options modified and added | Multiple choice |
|  | Preferred attributes of contraception in general | 1 | 11 | Ad-hoc translated, adapted | Multiple choice |
| **Subjective knowledge** | Self-assessment of familiarity with a particular contraceptive method | 15 | 4 | Ad-hoc translated, self-developed item added | 4-point Likert scale from 1 "I have never heard of it" to 4 "I know exactly what it is and how it works" |
|  | Knowledge about the mode of action, side effects, interactions and maintenance of HC effects | 4 | 47 | Ad-hoc translated, adapted | 5-point Likert scale from 1 "does not apply at all" to 5 "fully applies" |
| **Information needs** | Assessment of information sources used regarding the current main contraceptive method | 1 | 46 | Adapted, response options added | Multiple choice |
|  | Identification of the most important information source for the current main contraceptive method | 1 | 46 | Adapted, response options added | Single choice |
|  | Received and desired information from main and other information sources about the current main contraceptive method | 2 | 14 | Ad-hoc translated, adapted, items added | Multiple choice |
|  | Need for information on contraceptive methods other than the one used | 4 | 14 | Ad-hoc translated, adapted, items added | 5-point Likert scale from 1 "does not apply at all" to 5 "fully applies" |
| **Contraceptive counseling** | Received and desired information from healthcare provider about the current main contraceptive method | 2 | 14 | Translated, adapted, items added | Multiple choice |
|  | *Person Centered Contraceptive Counseling Scale (PCCC)*: Assessment of patient-centeredness in contraceptive counseling | 4 | 31,36 | Validated scale, translated via team translation protocol (TRAPD) | 5-point Likert scale from 1 "poor" to 5 "excellent" |
|  | *CollaboRATE*: Uptake of shared decision-making (in the clinical encounter from the patients' perspective) | 3 | 37,38, 40 | Validated German scale, adapted | 10-point Likert scale from 0 "no effort was made" to 9 "every effort was made" |
|  | *Control Preference Scale (CPS)*: Preferred and experienced degree of control of individuals regarding medical decisions | 2 | 40,41, 43,44 | Adapted German scale | 5-point Likert scale from 1 "only myself" to 5 "only my physician" |

The final survey comprised a total of 42 questions on 16 pages (between 1 and 11 questions per page) and could be completed in 15 to 20 minutes (see S2 Appendix for the English version of the survey and S3 Appendix for the German version of the survey). Some questions were only displayed if certain answer options of previous questions were selected. The survey was tested for comprehensiveness, ease of completion and accuracy regarding the time needed for completion by research assistants of the research group. The items were not randomized. The initial items contained demographic data including age, gender, place of birth, current place of residence, marital status, as well as information on (previous) pregnancies and number of children. Subsequently, participants were asked about their contraceptive behavior, attitudes, subjective knowledge and information needs. If participants had contraceptive counseling with their physician (gynecologist or other kind of physician) within the past 12 months, they were asked about their experience with the last visit that involved contraceptive counseling. For the purpose of this study, contraceptive counseling was defined as an active conversation between a physician, such as a gynecologist, and the individual regarding contraceptive options. If they had no consultation within the last 12 months, participants were redirected to the last page of the survey.

### Data analysis

Data were analyzed quantitatively via the statistical software SPSS version 29 (48). All questions were mandatory. Only participants who provided informed consent and were not excluded based on the screening questions were included in the analysis. Respondents who dropped out of the survey were generally retained in the dataset if they completed the relevant sections for a particular analysis. However, cases in which respondents only partially answered an item block (e.g., a scale with multiple items measuring one construct) were excluded from analysis regarding this item block to avoid bias in mean calculations and statistical analyses. Not all items contained in the questionnaire were evaluated as part of the analysis within this study. A complete overview of all items surveyed can be found in S2 and S3 Appendix. A descriptive presentation of all survey items can be found in S4 Appendix.

To compare HC users and non-HC users, participants were categorized into two groups defined by following criteria: the HC group included all participants who were using at least one hormonal contraceptive method (e.g., OCP, hormonal IUD, vaginal ring) at the time of answering the survey, while the non-HC group included all participants who were using at least one non-hormonal contraceptive method listed (condom, copper IUD, copper chain/ball, and natural contraceptive methods). If participants reported using both, a hormonal and non-hormonal contraceptive method, they were assigned to the HC group. Participants who stated “I do not use contraception” were not assigned to any group and were not included in group comparison. If participants indicated using “Other contraceptive methods”, the decision on group assignment was made based on the contraceptive indicated and via discussions within the study team. For group comparison analyses, only participants who completed the entire questionnaire were included.

Data were first analyzed descriptively. This included calculating means, standard deviations, and frequency distributions. To conduct the Student’s t-Test for the two independent groups (HC group and non-HC group), the Shapiro-Wilk test was used first to verify normal distribution of the groups. If the p-value of this test was greater than the significance level (α) of 5%, data in both groups were described as normally distributed. In this case, a test for variance homogeneity was performed using the Levene’s test. If the p-value from Levene’s test was greater than α (5%), this indicated that the variances were equal, and a t-Test for independent groups was performed.

## RESULTS

### Description of the data set

From May 10th to June 15th 2024, a total of 148 participants accessed the online survey. Of these, 14.9% (*n*=22) had to be excluded because they closed the survey after the introduction page (9.1%, *n*=2), refused to give informed consent (4.5%, *n*=1) or did not meet inclusion criteria (86.4%, *n*=19). 126 participants were included in data analysis and 105 participants fully completed the survey.

The 126 participants had an average age of 24.12 years (*SD*=4.86), most identified themselves as female (96.8%, *n*=122), were in a stable relationship (61.1%, *n*=77), and did not feel they belonged to any religion (44.4%, *n*=56). *N*=11 (8.7%) participants had been pregnant before. Most of the participants were born in Germany (96.0%, *n*=121), have a German citizenship (97.6%, *n*=123) and were residing in Thuringia (26.2%, *n*=33). For details on demographic data of participants, see Table 2.

**Table 2.**
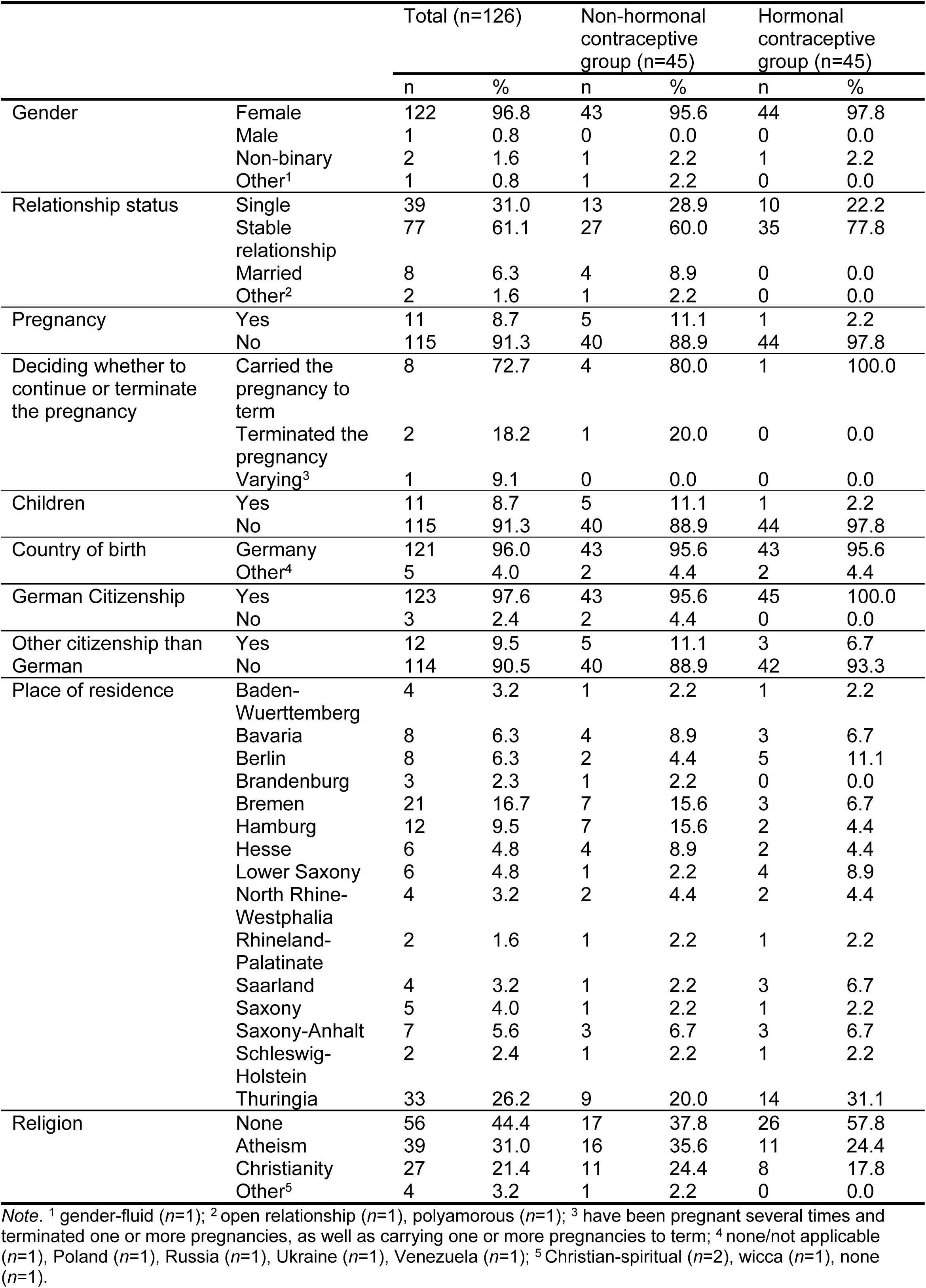
Demographic characteristics of all participants (*n*=126), hormonal (*n*=45) and non-hormonal (*n*=45) contraceptive group.

| | | Total ($n=126$) | | Non-hormonal contraceptive group ($n=45$) | | Hormonal contraceptive group ($n=45$) | |
| --- | --- | --- | --- | --- | --- | --- | --- |
|  |  | n | % | n | % | n | % |
| Gender | Female | 122 | 96.8 | 43 | 95.6 | 44 | 97.8 |
|  | Male | 1 | 0.8 | 0 | 0.0 | 0 | 0.0 |
|  | Non-binary | 2 | 1.6 | 1 | 2.2 | 1 | 2.2 |
|  | Other<sup>1</sup> | 1 | 0.8 | 1 | 2.2 | 0 | 0.0 |
| Relationship status | Single | 39 | 31.0 | 13 | 28.9 | 10 | 22.2 |
|  | Stable relationship | 77 | 61.1 | 27 | 60.0 | 35 | 77.8 |
|  | Married | 8 | 6.3 | 4 | 8.9 | 0 | 0.0 |
|  | Other<sup>2</sup> | 2 | 1.6 | 1 | 2.2 | 0 | 0.0 |
| Pregnancy | Yes | 11 | 8.7 | 5 | 11.1 | 1 | 2.2 |
|  | No | 115 | 91.3 | 40 | 88.9 | 44 | 97.8 |
| Deciding whether to continue or terminate the pregnancy | Carried the pregnancy to term | 8 | 72.7 | 4 | 80.0 | 1 | 100.0 |
|  | Terminated the pregnancy | 2 | 18.2 | 1 | 20.0 | 0 | 0.0 |
|  | Varying<sup>3</sup> | 1 | 9.1 | 0 | 0.0 | 0 | 0.0 |
| Children | Yes | 11 | 8.7 | 5 | 11.1 | 1 | 2.2 |
|  | No | 115 | 91.3 | 40 | 88.9 | 44 | 97.8 |
| Country of birth | Germany | 121 | 96.0 | 43 | 95.6 | 43 | 95.6 |
|  | Other<sup>4</sup> | 5 | 4.0 | 2 | 4.4 | 2 | 4.4 |
| German Citizenship | Yes | 123 | 97.6 | 43 | 95.6 | 45 | 100.0 |
|  | No | 3 | 2.4 | 2 | 4.4 | 0 | 0.0 |
| Other citizenship than German | Yes | 12 | 9.5 | 5 | 11.1 | 3 | 6.7 |
|  | No | 114 | 90.5 | 40 | 88.9 | 42 | 93.3 |
| Place of residence | Baden-Wuerttemberg | 4 | 3.2 | 1 | 2.2 | 1 | 2.2 |
|  | Bavaria | 8 | 6.3 | 4 | 8.9 | 3 | 6.7 |
|  | Berlin | 8 | 6.3 | 2 | 4.4 | 5 | 11.1 |
|  | Brandenburg | 3 | 2.3 | 1 | 2.2 | 0 | 0.0 |
|  | Bremen | 21 | 16.7 | 7 | 15.6 | 3 | 6.7 |
|  | Hamburg | 12 | 9.5 | 7 | 15.6 | 2 | 4.4 |
|  | Hesse | 6 | 4.8 | 4 | 8.9 | 2 | 4.4 |
|  | Lower Saxony | 6 | 4.8 | 1 | 2.2 | 4 | 8.9 |
|  | North Rhine-Westphalia | 4 | 3.2 | 2 | 4.4 | 2 | 4.4 |
|  | Rhineland-Palatinate | 2 | 1.6 | 1 | 2.2 | 1 | 2.2 |
|  | Saarland | 4 | 3.2 | 1 | 2.2 | 3 | 6.7 |
|  | Saxony | 5 | 4.0 | 1 | 2.2 | 1 | 2.2 |
|  | Saxony-Anhalt | 7 | 5.6 | 3 | 6.7 | 3 | 6.7 |
|  | Schleswig-Holstein | 2 | 2.4 | 1 | 2.2 | 1 | 2.2 |
|  | Thuringia | 33 | 26.2 | 9 | 20.0 | 14 | 31.1 |
| Religion | None | 56 | 44.4 | 17 | 37.8 | 26 | 57.8 |
|  | Atheism | 39 | 31.0 | 16 | 35.6 | 11 | 24.4 |
|  | Christianity | 27 | 21.4 | 11 | 24.4 | 8 | 17.8 |
|  | Other<sup>5</sup> | 4 | 3.2 | 1 | 2.2 | 0 | 0.0 |

*Note.* <sup>1</sup> gender-fluid ( $n=1$ ); <sup>2</sup> open relationship ( $n=1$ ), polyamorous ( $n=1$ ); <sup>3</sup> have been pregnant several times and terminated one or more pregnancies, as well as carrying one or more pregnancies to term; <sup>4</sup> none/not applicable ( $n=1$ ), Poland ( $n=1$ ), Russia ( $n=1$ ), Ukraine ( $n=1$ ), Venezuela ( $n=1$ ); <sup>5</sup> Christian-spiritual ( $n=2$ ), wicca ( $n=1$ ), none ( $n=1$ ).

### Contraceptive Use

83.5% (*n*=101) of the participants indicated currently using at least one contraceptive method. The three most frequently used methods were the male condom (52.5%; *n*=53), the combined oral contraceptive pill (23.8%; *n*=24), and the progestin-only pill (9.9%; n=10). Participants were asked to refer to the contraceptive method, they define as the most important or main method used at the time of the survey when answering the subsequent items. More than half of the participants using contraception at the time of the survey had been using it for more than three years (53.5%; *n*=53). Twenty participants (16.5%, *n*=20) stated not using any form of contraception, with the main reason being that they were not sexually active (45.0%, *n*=9). The overall satisfaction level with the contraceptive method used at the time of the survey was found to be high. 36.7% (*n*=44) expressed being very satisfied, while 48.3% (*n*=58) were quite satisfied with their current contraceptive method. Overall, 76.7% (*n*=92) stated that they would not or rather not change their current main method of contraception. Further details on contraceptive use characteristics of the participants can be found in Table 3.

**Table 3.** Overview of contraceptive use, duration, satisfaction, willingness to change, and reasons for non-use.

|  | *n* | % |
| --- | --- | --- |
| **Contraceptive method used ($n=101$)<sup>1</sup>** |  |  |
| Condom | 53 | 52.5 |
| Combined contraceptive pill | 24 | 23.8 |
| Progestin-only pill | 10 | 9.9 |
| Natural contraceptive method | 9 | 8.9 |
| Copper intrauterine device | 9 | 8.9 |
| Hormonal intrauterine device | 6 | 5.9 |
| Pill, but not sure which one | 5 | 5.0 |
| Copper chain/copper ball | 4 | 4.0 |
| Vaginal ring | 3 | 3.0 |
| Other<sup>2</sup> | 4 | 2.0 |
| **Duration of use of current main method ($n=99$)<sup>3</sup>** |  |  |
| Less than one month | 1 | 1.0 |
| Between one and six months | 4 | 4.0 |
| Between seven and twelve months | 16 | 16.2 |
| Between 13 months and three years | 25 | 25.3 |
| More than three years | 53 | 53.5 |
| **Satisfaction with current main method ($n=120$)** |  |  |
| Very satisfied | 44 | 36.7 |
| Rather satisfied | 58 | 48.3 |
| Neither | 13 | 10.8 |
| Less satisfied | 5 | 4.2 |
| **Willingness to change current main method ($n=120$)** |  |  |
| No | 57 | 47.5 |
| Rather no | 35 | 29.2 |
| Rather yes | 21 | 17.5 |
| Yes | 7 | 5.8 |
| **Reasons for not using contraception (n=20)<sup>1</sup>** |  |  |
| I am sexually not active | 9 | 45.0 |
| I would like to get pregnant | 1 | 5.0 |
| I do not want any side effects | 2 | 10.0 |
| Other<sup>4</sup> | 8 | 40.0 |

*Note.* Data excluding participants not using contraceptive methods, varied subsample sizes per item, <sup>1</sup> multiple answers were possible. <sup>2</sup> sterilization of the partner ( $n=2$ ), contraceptive patch ( $n=1$ ), withdrawal ( $n=1$ ), <sup>3</sup> Two participants (1.65%) stated that they were not using contraception and chose a method at the same time. Therefore, they were not forwarded to the question about the duration of the current main contraceptive method, but were able to answer subsequent questions, such as satisfaction with the respective contraceptive method, <sup>4</sup> current pregnancy ( $n=2$ ), homosexual ( $n=6$ ).

### Group comparison of HC users and non-HC users

Of *n*=105 participants, who were included in the group comparison analysis because they fully completed the survey, 14.3% (*n*=15) did not use contraception and were therefore also excluded from the comparison of HC users and non-HC users. According to the above-described criteria, 45 participants could be assigned to the “HC group”, while 45 participants could be assigned to the “non-HC group”. A case-by-case decision on group assignment via discussions within the study team had to be made for 3 participants (2.8%). According to this discussion, one participant was not assigned to any group and was considered to be not using contraception. Due to the group size of 45 participants each, the calculated sample size was not reached. Consequently, a post-hoc analysis was conducted using G*Power for the actual group size achieved for two independent groups. The statistical power calculated was 0.76, which means that the calculations had a 76% probability of detecting an actual effect if it existed. A subsequent sensitivity analysis showed a Cohen’s d value of 0.69, indicating a medium to large effect. To control alpha error type-I, a Bonferroni correction was applied, adjusting the alpha level to 0.0167. Statistical significance for following group differences were assessed using this corrected alpha level.

The HC group was slightly younger, with an average age of 22.84 years (*SD*=3.59), compared to the non-HC group, which had an average age of 25.40 years (*SD*=5.62). This difference was statistically significant (t(88)=-2.57, *p*=.012).

### Attitude towards hormonal contraception

The average attitude towards HC was found to be slightly negative, with a mean score of 3.28 (*SD*=0.70) on a scale from 1 (‘positive attitude towards HC’) to 5 (‘negative attitude towards HC’). When comparing groups, the mean attitude score for non-HC users (*M*=3.67, *SD*=0.48) was higher than that of HC users (*M*=2.78, *SD*=0.54) (see Table 4 for single item mean scores). The difference in attitudes towards HC between the two groups was statistically significant (*t*(88)=-8.37, *p*<.001). The majority of participants indicated that they believe hormones used in contraception can cause hormonal disorders (82.5%; *n*=94) and/or thrombosis (76.3%; *n*=87). When asked about the individual relevance of different contraceptive attributes, all respondents indicated that prevention of pregnancy is most important for them (100.0%; *n*=114), followed by experiencing few side effects (96.5%; *n*=110), and good tolerance (94.7%; *n*=108).

**Table 4.**
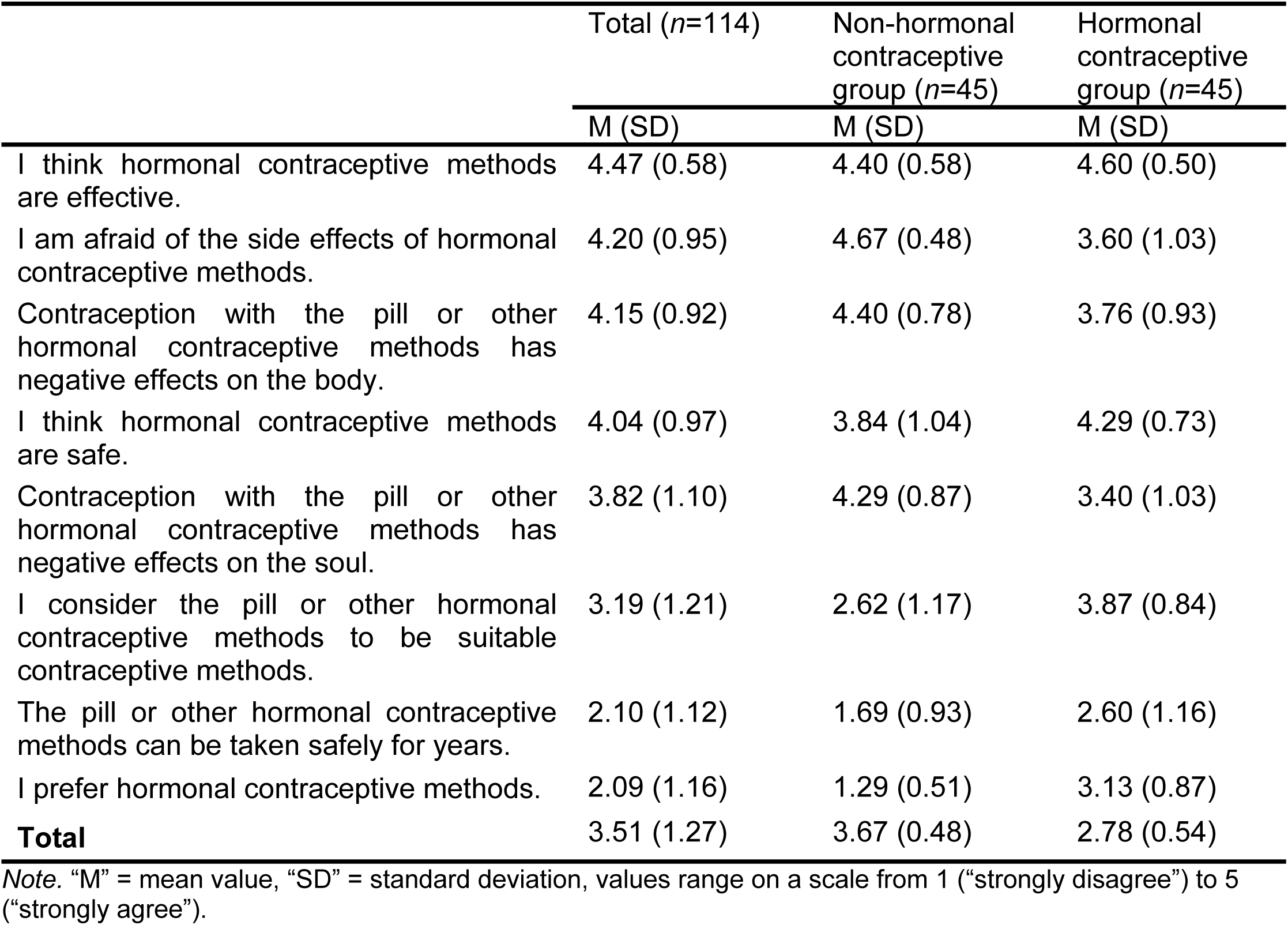
Mean values and standard deviations for items regarding the attitude towards hormonal contraception.

### Subjective knowledge of contraceptive methods

Only 6.3% of the participants (*n*=7) reported having good or basic knowledge of all 15 methods presented (e.g., rated as a 1 or 2 on a scale from 1 “I know exactly what it is and how it works” to 4 “I have never heard of it”). On average, participants had good or basic knowledge of nine methods (see Fig 1).

**Fig 1.**
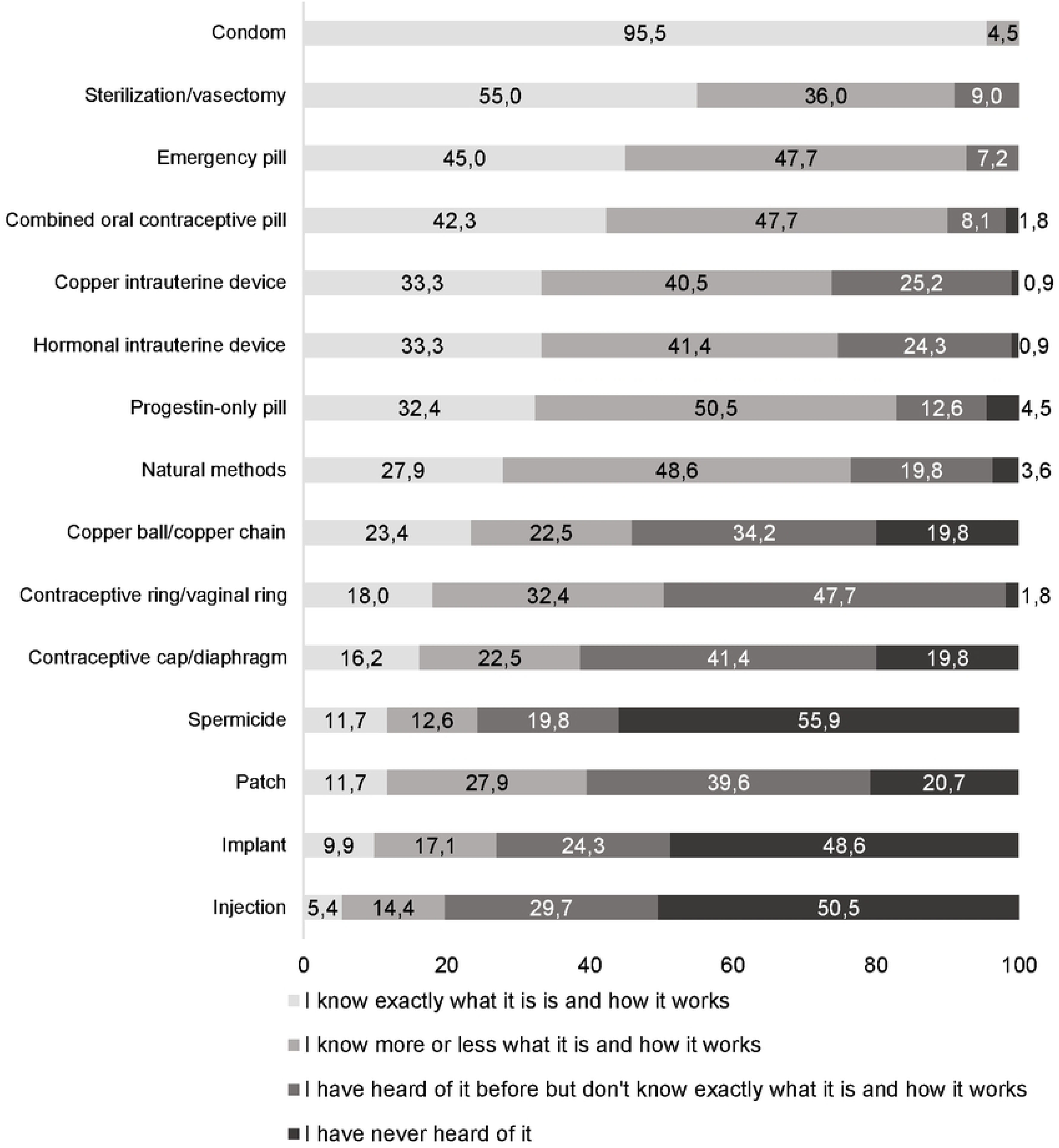
Relative frequency for subjective knowledge of the different contraceptive methods (n=111)

Comparing groups, subjective knowledge was generally lower for the majority of methods in the HC group (see S5 Appendix). However, no statistically significant difference in knowledge was found between the two groups (*t*(88)=-0.89, *p*=.379).

### Information needs regarding contraceptive methods

On average, participants reported using three sources of information (*M*=2.70, *SD*=1.39). The gynecologist was identified as the most frequently used source (78.2%, *n*=86), followed by friends (50.0%, *n*=55) (Table 5). The three sources that were least frequently used were the radio (*n*=0), sex films (*n*=2) and online-chat with others (*n*=4).

**Table 5.**
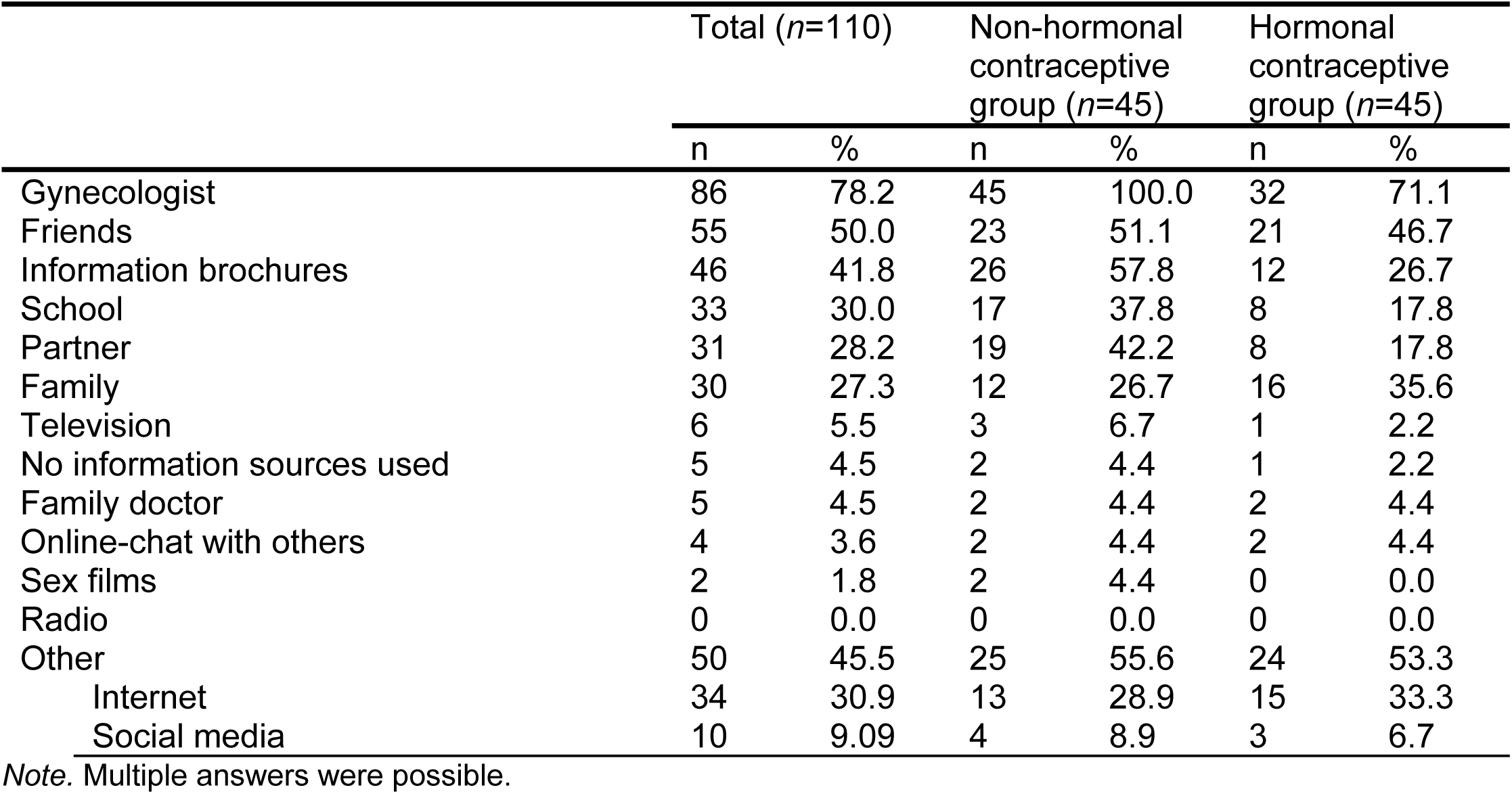
Absolute and relative frequencies for the information sources used for the current main contraceptive method.

| | Total ($n=110$) | | Non-hormonal contraceptive group ($n=45$) | | Hormonal contraceptive group ($n=45$) | |
| --- | --- | --- | --- | --- | --- | --- |
|  | n | % | n | % | n | % |
| Gynecologist | 86 | 78.2 | 45 | 100.0 | 32 | 71.1 |
| Friends | 55 | 50.0 | 23 | 51.1 | 21 | 46.7 |
| Information brochures | 46 | 41.8 | 26 | 57.8 | 12 | 26.7 |
| School | 33 | 30.0 | 17 | 37.8 | 8 | 17.8 |
| Partner | 31 | 28.2 | 19 | 42.2 | 8 | 17.8 |
| Family | 30 | 27.3 | 12 | 26.7 | 16 | 35.6 |
| Television | 6 | 5.5 | 3 | 6.7 | 1 | 2.2 |
| No information sources used | 5 | 4.5 | 2 | 4.4 | 1 | 2.2 |
| Family doctor | 5 | 4.5 | 2 | 4.4 | 2 | 4.4 |
| Online-chat with others | 4 | 3.6 | 2 | 4.4 | 2 | 4.4 |
| Sex films | 2 | 1.8 | 2 | 4.4 | 0 | 0.0 |
| Radio | 0 | 0.0 | 0 | 0.0 | 0 | 0.0 |
| Other | 50 | 45.5 | 25 | 55.6 | 24 | 53.3 |
| Internet | 34 | 30.9 | 13 | 28.9 | 15 | 33.3 |
| Social media | 10 | 9.09 | 4 | 8.9 | 3 | 6.7 |

Note. Multiple answers were possible.

When asked which of those information sources were most important for them, 36.7% (*n*=40) out of 109 participants indicated their gynecologist as the most important source, 30.3% (*n*=33) named others, with the internet being the most frequently cited, and 14.7% (*n*=16) identified friends as the most important source of information. The information participants received most from their most important source of information included the effectiveness/safety of contraceptive methods (69.7%; *n*=76), their mode of action/function (68.8%; *n*=75) and instructions for use (66.1%; *n*=72). Most information received from other than the most important source was reports from others (56.9%, *n*=62), side effects (e.g., changes in the menstrual cycle; 41.3%, *n*=45) and effectiveness/safety (35.8%, *n*=39). The highest need for information of all participants was related to side effects (41.3%; *n*=45) and interactions (33.0%; *n*=36). However, 37.6% (*n*=41) of participants indicated that they do not need any further information. Participants were further asked to indicate, which information they need on other contraceptive methods apart from the main one they used at the time of the survey. The highest need for information regarding contraceptive methods other than the current main contraceptive method was regarding side effects (e.g., changes in the menstrual cycle; M=3.88, SD=1.11), followed by interactions with other contraceptive methods (e.g., with other medications or alcohol; M=3.65, SD=1.17), mechanism of action (M=3.51, SD=1.16), and instructions for use (M=3.36, SD=1.18) (scale from 1 (“does not apply at all”) to 5 (“fully applies”)). Furthermore, a higher need for information was observed among participants who were less satisfied with their current main contraceptive method. More details on information sources and needs of participants can be found in S5 Appendix.

### Evaluation of contraceptive counseling

45 participants indicated that they received contraceptive counseling within the 12 months prior to study participation. Of these, 28 were HC users, while 14 participants were non-HC users. Nearly all of them (97.8%; *n*=44) were counseled by a gynecologist, with only *n*=1 (2.2%) receiving counseling from a family doctor. Within the counseling session, most of the participants received information about their current contraceptive method regarding side effects (e.g., changes in the menstrual cycle; 44.4%, *n*=20), the mode of action/function (46.7%, *n*=20), and the effectiveness/safety (44.4%, *n*=20) (see Fig 2). Almost half of the participants did not require any further information on their current main contraceptive method (46.7%, *n*=21). More information was needed on interactions (e.g., with other medications or alcohol; 35.6%, *n*=16), side effects (e.g., changes in the menstrual cycle; 33.4%, *n*=15) and mode of action/function (26.7%, *n*=12).

**Fig 2.**
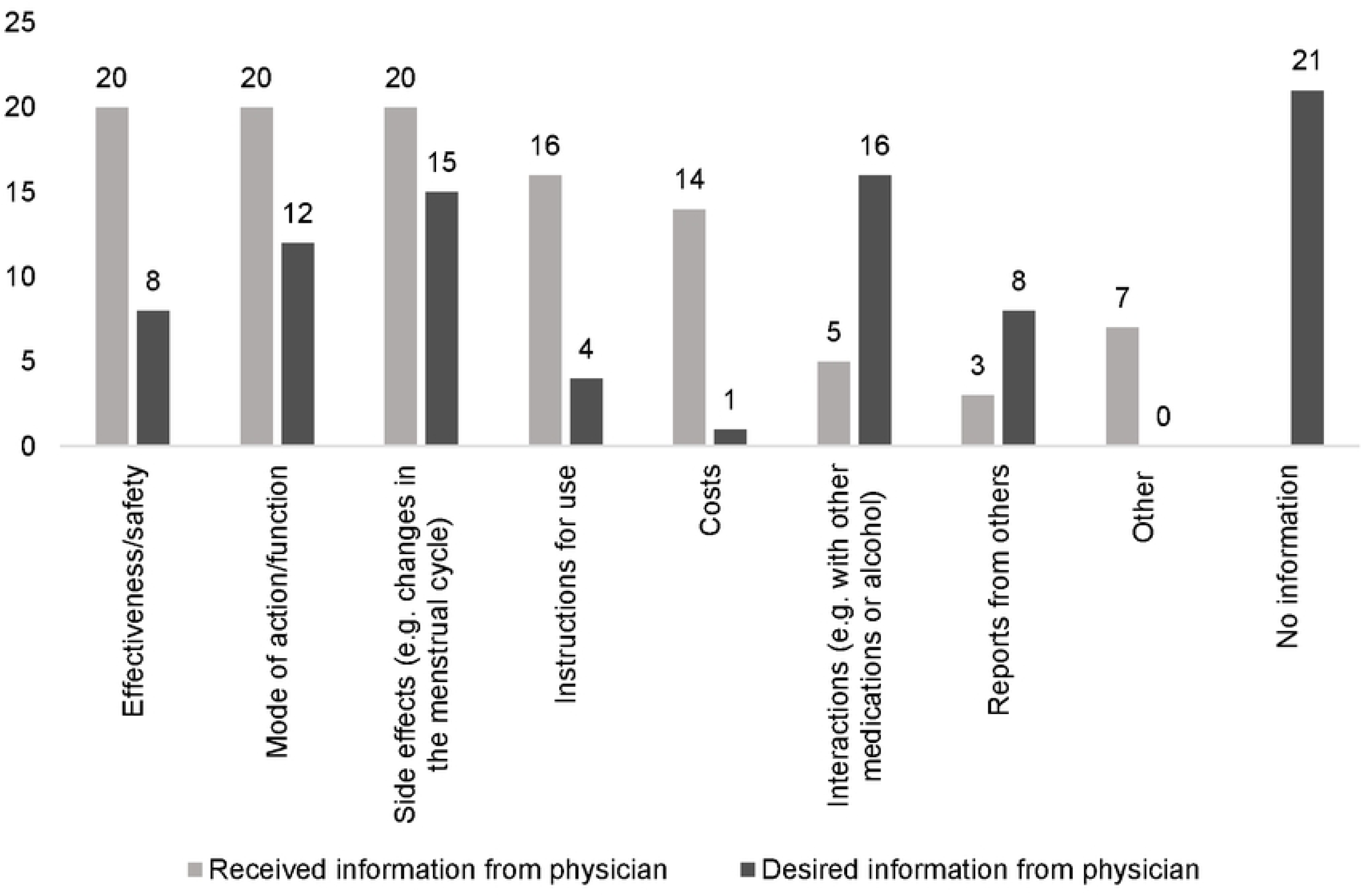
Absolute frequency of received and desired information within the last contraceptive counseling session (*n*=45) *Notes*. Multiple answers were possible. “No information” was not only a response option for the category “Received information from healthcare provider”. ^1^ none/not applicable (*n*=2), switching contraceptives (*n*=2), mechanism of action regarding endometriosis (*n*=1), general information/support (*n*=1), alternative contraceptives (*n*=1)

Additionally, nearly half of the participants (46.7%, *n*=21; HC-group: 35.6%, *n*=16; non-HC group: 11.1%, *n*=5) wanted to receive additional information regarding non-HC, while only 16.7% (*n*=7; HC-group: 13.3%, *n*=6; non-HC-group: 2.2%, *n*=1) wished for further details about HC (see S5 Appendix). 44.4% (*n*=20; HC-group: 24.4%, *n*=11; non-HC-group: 20.0%, *n*=9) had no need for more information from their physician regarding their current contraceptive method. In case that further information was desired, the majority of participants sought most information on effectiveness/safety (HC: 71.4%; *n*=5; non-HC: 75.0%; *n*=18), side effects (e.g., changes in the menstrual cycle; HC: 100.0%; n=7; non-HC: 75.0%; *n*=18), mode of action (HC: 85.7%; *n*=6; non-HC: 70.8%; *n*=17) and instructions for use (HC: 71.4%; *n*=5; non-HC: 70.8%; n=17). Overall, the participants tended to be less satisfied with the information provided by the physician regarding their current main contraceptive method, with a mean score of 2.51 (SD=1.22) on a scale from 1 (“not satisfied”) to 5 (“very satisfied”) (see Table 6).

**Table 6.**
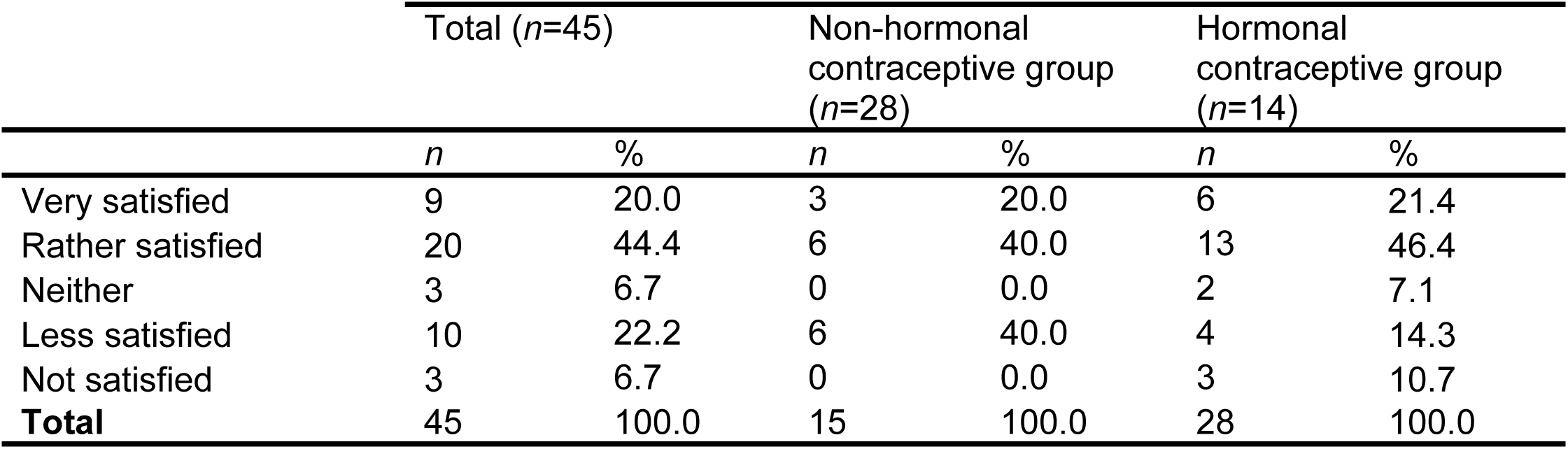
Absolute and relative frequencies for satisfaction with the information received from the physician.

| | Total ($n=45$) | | Non-hormonal contraceptive group ($n=28$) | | Hormonal contraceptive group ($n=14$) | |
| --- | --- | --- | --- | --- | --- | --- |
| | $n$ | % | $n$ | % | $n$ | % |
| Very satisfied | 9 | 20.0 | 3 | 20.0 | 6 | 21.4 |
| Rather satisfied | 20 | 44.4 | 6 | 40.0 | 13 | 46.4 |
| Neither | 3 | 6.7 | 0 | 0.0 | 2 | 7.1 |
| Less satisfied | 10 | 22.2 | 6 | 40.0 | 4 | 14.3 |
| Not satisfied | 3 | 6.7 | 0 | 0.0 | 3 | 10.7 |
| **Total** | **45** | **100.0** | **15** | **100.0** | **28** | **100.0** |

The overall PCCC (31,36) mean score for the 45 participants, who had received contraceptive counseling within the past 12 months, was 3.48 (*SD*=1.22; non-HC-group: *M*=3.30, *SD*=1.25; HC-group: *M*=3.61, *SD*=1.63) on a scale from 1 (“poor”) to 5 (“excellent”) (see Fig 3).

**Fig 3.**
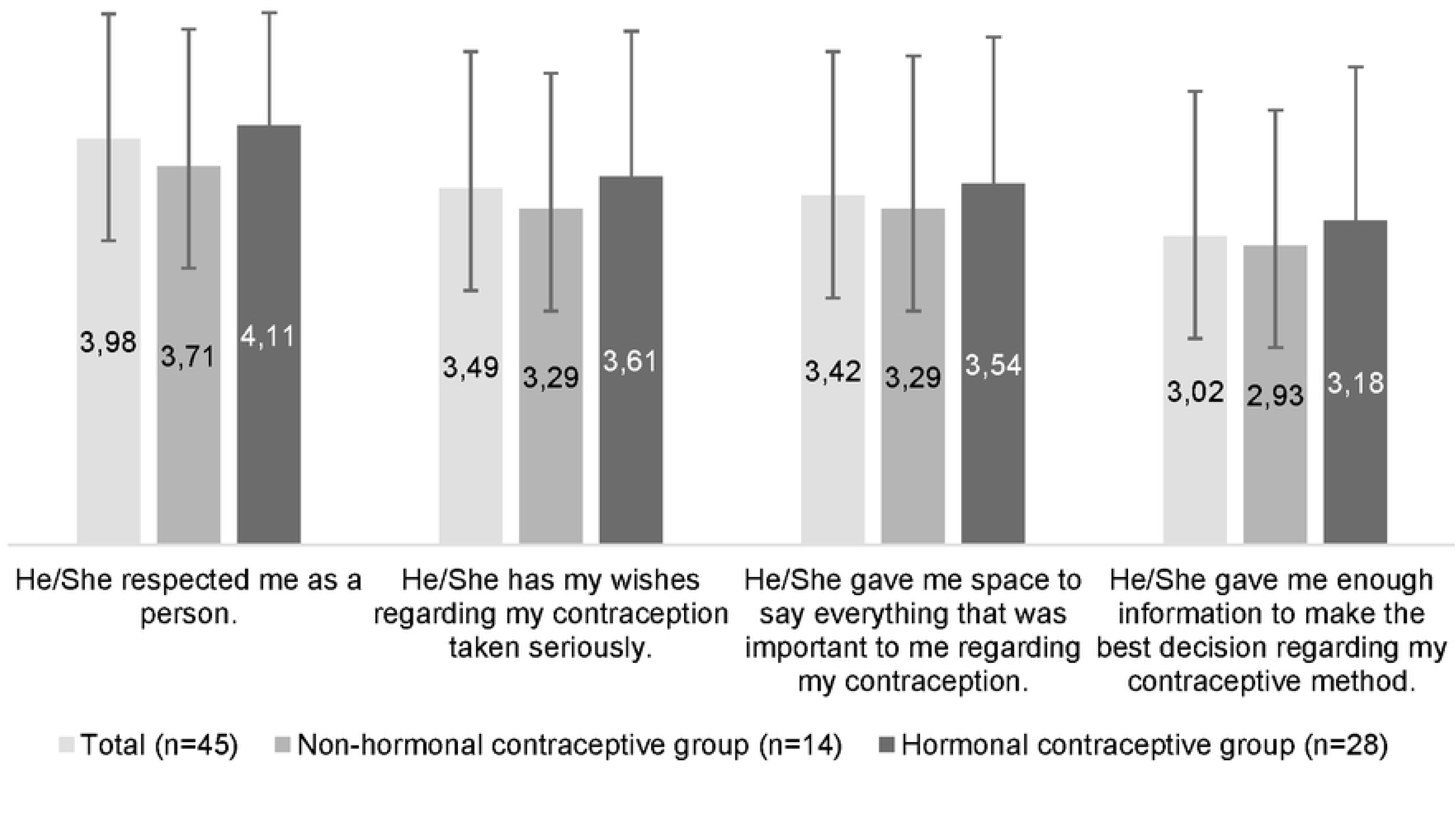
Mean values and standard deviations for items of the Person-Centered Contraceptive Counseling (31,36) questionnaire *Note*. On a scale from 1 “poor” to 5 “excellent”.

The mean score for the degree of SDM uptake from the patients’ perspective (CollaboRATE scale) was 5.81 (*SD*=2.60; non-HC-group: *M*=5.76, *SD*=2.47; HC-group: *M*=6.07, *SD*=2.88) on a scale from 0 (“no effort was made”) to 9 (“every effort was made”) (41). The mean values of the individual items on the scale were comparable (see Fig 4).

**Fig 4.**
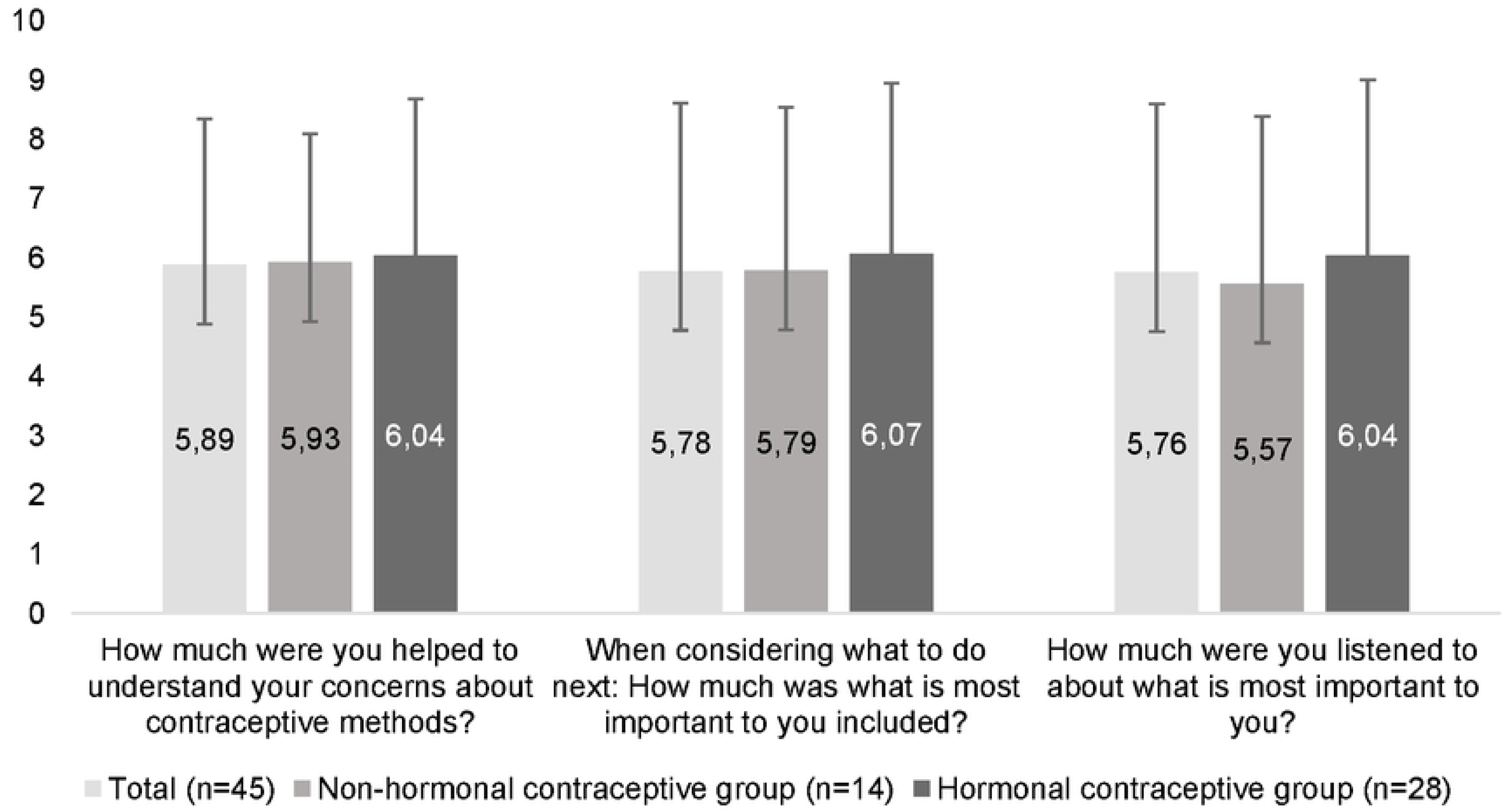
Mean values and standard deviations for items of the CollaboRATE scale (37–39) *Note*. On a scale from 0 “no effort was made” to 9 “every effort was made”.

As part of an exploratory analysis, correlations between the general satisfaction with the current contraceptive method and the PCCC as well as CollaboRATE mean values were calculated (see Table 7). Participants who expressed high satisfaction with their current contraceptive method reported higher mean scores on both scales. In addition, HC users and non-HC users were compared regarding their values on the PCCC scale and CollaboRATE scale. Thereby, n=28 HC user and n=14 non-HC user could be included in analysis via t-test. However, no statistically significant differences were observed between the two groups on either scale (PCCC: *t*(40)=0.74, *p*=0.47, CollaboRATE: *t*(40)=0.33, *p*=0.74).

**Table 7.**
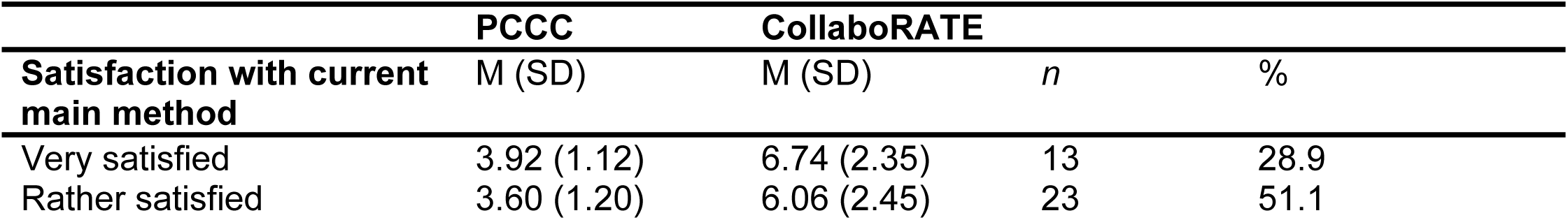
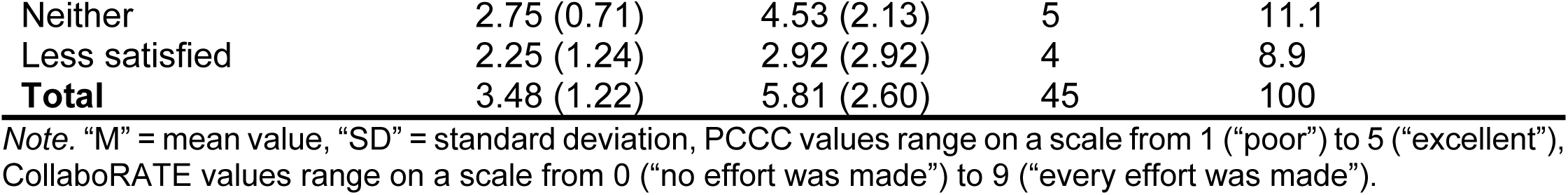
Person-Centered Contraceptive Counseling(30,35) and CollaboRATE scale (37,38,40) in relation to satisfaction.

|  | PCCC | CollaboRATE |  |  |
| --- | --- | --- | --- | --- |
| Satisfaction with current main method | M (SD) | M (SD) | *n* | % |
| Very satisfied | 3.92 (1.12) | 6.74 (2.35) | 13 | 28.9 |
| Rather satisfied | 3.60 (1.20) | 6.06 (2.45) | 23 | 51.1 |
| Neither | 2.75 (0.71) | 4.53 (2.13) | 5 | 11.1 |
| Less satisfied | 2.25 (1.24) | 2.92 (2.92) | 4 | 8.9 |
| **Total** | **3.48 (1.22)** | **5.81 (2.60)** | **45** | **100** |

Note. "M" = mean value, "SD" = standard deviation, PCCC values range on a scale from 1 ("poor") to 5 ("excellent"), CollaboRATE values range on a scale from 0 ("no effort was made") to 9 ("every effort was made").

Regarding decision-making about contraceptive methods, 44.4% (*n*=20) of participants who had received a counseling session within the past 12 months stated that the decision should be made entirely on their own (Control Preference Scale (41), see Fig 5). *N*=17 participants (37.8%) experienced that they could decide about the method used alone. No participant indicated that the decision should mainly be made by the physician, although 20.0% (*n*=9) reported having experienced this.

**Fig 5.**
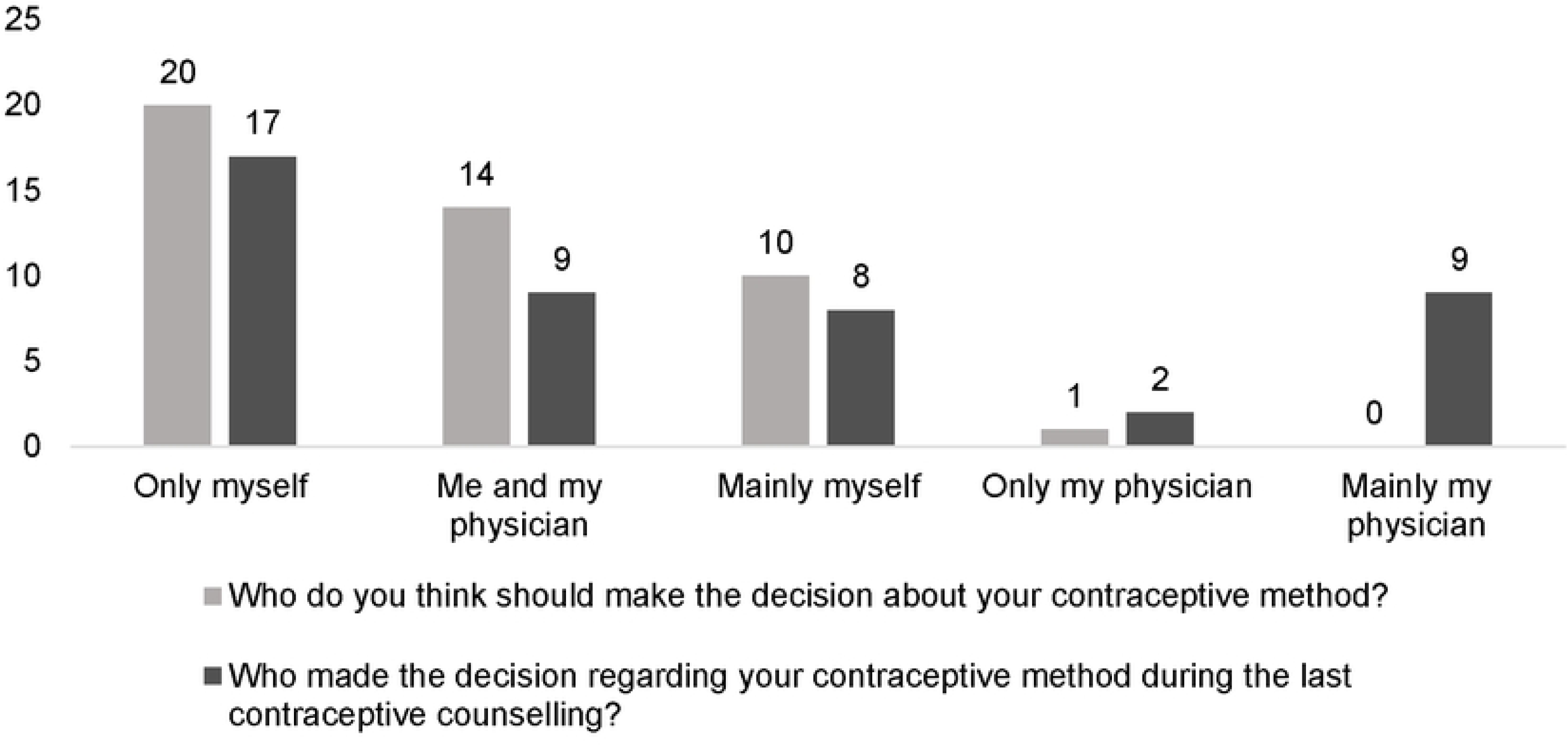
Absolute frequency of the Control Preference Scale (41) regarding the last contraceptive counseling session (*n*=45)

## DISCUSSION

This study provides insights into contraceptive use, attitudes towards HC, subjective knowledge about contraceptive methods, information needs and decision-making processes among 126 psychology students in Germany. A high prevalence of contraceptive use was observed among all participants, with the condom, combined oral contraceptive pill and progestin-only pill being the most commonly used methods. Differences were found between HC users and non-HC users, particularly regarding age, satisfaction with the current main contraceptive method, attitudes towards HC, knowledge of the various contraceptive methods and need for information regarding contraception. Results showed a limited uptake of PC and SDM in contraceptive counseling. During contraceptive counseling, the majority of participants made the treatment decision either independently or in collaboration with their physician. In some cases, the decision was made exclusively by the physician. However, none of the participants favored exclusive provider-led decision-making. Some participants felt that they were not provided with sufficient information to make an informed decision regarding their contraceptive method, indicating potential for improvement in patient-centered care and SDM to ensure that counseling sessions address individual concerns and preferences, with the aim of achieving higher satisfaction and better-informed decisions.

The findings of this study are consistent with others that have investigated people’s contraceptive behavior, attitudes, knowledge, information needs and counseling experience, both nationally and internationally (4,7). Our findings regarding use of and satisfaction with the contraceptive methods are congruent with results of other studies (4,46,47). Similar to studies from Saudi Arabia and Germany, participants in our study described hormonal contraceptive methods as harmful to the body and were afraid of the side effects (46,47). Nevertheless, the non-HC users in our study reported higher satisfaction, which may be because their current contraceptive method matches their preferred method. Furthermore, effectiveness and safety was also rated as the most important attribute of contraceptive methods in several other studies (4,19). Gaps in knowledge were also described in other studies conducted in Europe and the USA (4,49). Only a small proportion of participants in this study reported good or basic knowledge of the most popular contraceptive methods, which is consistent to the findings of the TANCO study, which conducted unmet needs in contraceptive counseling and choices of 6.027 women in European countries (4). Sharma et al. (2021) surveyed 130 participants in the US regarding their knowledge of contraceptive methods (particularly IUDs), whereas many reported having heard of IUDs at least once. However, it was observed that the details of participants’ knowledge about the use of IUDs were lacking (49).

In contrast, the present study indicates a high level of knowledge regarding the maintenance of the effectiveness of hormonal contraceptive methods, including the hormonal IUD. The use of information sources in this study is also reflected in results of the survey on contraceptive behavior conducted by the Federal Center for Health Education (german: Bundeszentrale für gesundheitliche Aufklärung) in 2023 (7). Similarly, gynecologists were named as the most frequently used source of information (7). The quality of contraceptive counseling was reported as good as in the TANCO and a study by Bitzer et al. (2021) (4,19). Concurrently, the present study demonstrated a strong demand for further information on side effects (e.g., changes in the menstrual cycle) and interactions (e.g., with other medications or alcohol). This finding indicates that current counseling practice may not adequately meet patients’ information needs. Similar results were obtained in the TANCO study, where 60% of participants indicated an interest in receiving more information from their HCP (4). It was also reported that HCPs tended to underestimate participants’ interest in receiving more information, as well as their interest in non- or low-hormonal contraceptive methods (4). This is congruent with the participants’ satisfaction with the information received from their physician in the study at hand: more than half of our participants were less or not at all satisfied with the information provided by their physician. In contrast, a study from the USA on postpartum contraceptive needs reported that over 85% of participants rated the information they received about contraceptive methods and the discussion about them as acceptable (50). These results do not reflect those of our study, in which the majority of participants reported having made the decision on their own or with their physician. In summary, although some of the needs of German psychology students for contraceptive counseling and decision-making are already covered, there are still gaps between desired and perceived PC in contraceptive counseling in Germany. PC and SDM in contraceptive counseling in Germany has to improve to meet to criteria for high quality contraceptive counseling which focus on people’s preferences and concerns as recommended by the World Health Organization (WHO) (30).

### Strengths and Limitations

A strength of this study is its direct comparison between HC users and non-HC users. This provides insight into different perceptions of contraceptive use, attitudes towards HC and general subjective knowledge about HC. By applying an a priori planned German-wide recruitment strategy, it was possible to collect data from participants in almost every federal state. The study is strengthened by the inclusion of participants of a wide age range using various contraceptive methods, which emphasizes the heterogeneity of this group. On the other hand, defining psychology students as the target group has to be discussed as a limitation, as this group is not a representative sample of the German population. Moreover, the findings are culturally and regionally specific, as this study only included students at German universities and institutions of higher education and therefore cannot be transferred to other countries and cultures or to German population groups with different education or socio-economic status. In future studies, a broader population with a diverse range of education levels and socio-economic status would be beneficial in order to better contextualize the results and evaluate those factors regarding their influence on contraceptive use, attitudes towards HC, knowledge and information needs. In addition, the translation process of several survey items may represent a limitation of this study. Single items, which were adapted from other studies, were neither discussed with the target group nor tested in cognitive interviews. This may have led to variations in interpretation due to cultural and linguistic differences (51). However, survey items from validated scales were translated via a structured team translation protocol. Another limitation was the small sample size. 29.05% of all participants who accessed the survey did not complete it. The main reason for this is presumed to be the absence of an incentive. The sample size of 126 participants that were included in the analysis limits the generalizability of the results to the target population. Furthermore, the study provides insight at a certain point in the respondents’ lives. Longitudinal studies could help to gain deeper insight on changes of satisfaction and discontinuation of contraceptive methods, attitudes, knowledge and information needs over time (52). Given the variety of contraceptive methods available, which differ in characteristics, side effects, cost and availability, preferences and needs may change over the life course (52).

## CONCLUSION

This study provides new insights into the attitudes, towards HC subjective knowledge, information needs and quality of PC and SDM in contraceptive counseling among psychology students in Germany. Our findings of unmet information needs underline the need for improved, participatory and need-oriented contraceptive counseling. Future research should include more diverse populations, use qualitative methods to examine individual experiences and needs and implement appropriate approaches to strengthen SDM and PC. A lack of research has been observed with regard to the extent to which partners are involved in the decision-making process regarding contraceptive methods. The examination of cis-men and partners also represents a research gap that would be important to fill in order to gain a deeper understanding of their perspective. To meet contraceptive needs of individuals according to WHO recommendations, provision of comprehensive information about contraception, transparent information about all available options and taking into account individual circumstances is key to realize informed and SDM in contraceptive counseling.

## Data Availability

The data set underlying this research article is available from the OSF repository (DOI 10.17605/OSF.IO/KWUQN).

https://doi.org/10.17605/OSF.IO/KWUQN

## SUPPORTING INFORMATION

S1 Appendix: Checklist for reporting results of internet e-surveys (CHERRIES)

S2 Appendix: Online survey on hormonal contraceptive methods, translated English version

S3 Appendix: Online-survey on hormonal contraceptive methods, original German version

S4 Appendix: Items of the online survey and descriptive data of all items

S5 Appendix: Analysis of further research question-related items

## LIST OF ABBREVIATIONS

AL: Anja Lindig
CHERRIES: Checklist for Reporting Results of Internet E-Surveys
HC: Hormonal Contraception
HCP: Healthcare Provider
IUD: Intrauterine Device
JS: Johanna Seiwert
MR: Mareike Rutenkröger
OCP: Oral Contraceptive Pill
PC: Person-Centeredness
SDM: Shared Decision-Making
TANCO: Thinking About Needs in Contraception study
TRAPD: Translation, Review, Adjudication, Pretesting and Documentation
VL: Vanessa Le

## DECLARATIONS

## Acknowledgements

We sincerely thank the researchers at the Department of Medical Psychology, University Medical Centre Hamburg-Eppendorf, for proofreading the survey. We especially thank Vanessa Le and Mareike Rutenkröger for their support in translation of the PCCC. We furthermore thank our participants for their trust and for taking part in our study.

## Ethics approval and consent to participate

The study was carried out according to the latest version of the Helsinki Declaration of the World Medical Association. Principles of good scientific practice have been respected. The study was approved by the Psychological Ethics Committee of the Center for Psychosocial Medicine of the University Medical Center Hamburg-Eppendorf, Germany (LPEK-0771). Standards of research ethics were met. This includes that study participation was voluntary and no foreseeable risks for participants resulted from the participation. Participants were fully informed about the aims of the study, data collection, and the use of collected data beforehand in written form. Informed consent was sought prior to participation by clicking on the respective button in the online survey form beneath the study information. Preserving principles of data sensitivity, data protection and confidentiality requirements were met.

## Consent for publication

Not applicable

## Availability of data and materials

The datasets analyzed in this study are available from the OSF data repository of this study (DOI 10.17605/OSF.IO/KWUQN).

## Author contributions

The study was conceptualized and designed by AL and JS. Both authors were responsible for methodology. JS developed the survey with supervision of AL. AL was part of the team, which translated and adapted the PCCC via the TRAPD protocol. JS was responsible for participants’ recruitment and data analysis. The first draft of this manuscript was written by JS. AL was engaged in critical revisal of the manuscript for important intellectual content. All authors have reviewed the final manuscript, approved the published version, and agreed to take responsibility for the work.

## Checklist for reporting statement

This manuscript was prepared in accordance with the CHERRIES, which is appropriate for the design of this study (34). The aim of using the checklist was to ensure transparency, comprehensiveness and replicability of the study reports. A completed checklist is provided in the appendix (S1 Appendix), indicating the pages in the manuscript where each checklist item is addressed. Checklist items that were not applicable or not reported are explained and justified in the checklist.

